# Arkangel AI, OpenEvidence, ChatGPT, Medisearch: are they objectively up to medical standards? A real-life assessment of LLMs in healthcare

**DOI:** 10.1101/2025.09.23.25336206

**Authors:** Natalia Castano-Villegas, María Camila Villa, Katherine Monsalve, Isabella Llano, Laura Velásquez, José Zea

## Abstract

**Background:** Large language models (LLMs) are increasingly used in healthcare, but standardized benchmarks fail to capture their validity and safety in real-world scenarios. Evaluating their quality and reliability is critical for safe integration into practice.

**Methods:** Four fictitious clinical vignettes (orthopedics, pediatrics, gynecology, psychiatry) were developed by independent specialists and tested in four conversational agents: ArkangelAI, OpenEvidence, ChatGPT, and Medisearch. Each vignette included four questions (diagnosis, management, research, and general knowledge). Responses were evaluated by four external clinicians using an eight-criterion Likert scale: 1–2 = dissatisfaction, 3 = neutral, 4–5 = satisfaction, 6 = not applicable. The criteria considered correctness, consensus, bias, standard of care, updated information, patient safety, real sources in references, and context-awareness. Response times were measured with medians and interquartile ranges (IQR). Results were reported as frequencies. Hypothesis tests were applied (α = 0.05).

**Results:** We assessed 128 question–answer (Q&A) pairs (1024 evaluations). ArkangelAI-Deep was the highest in satisfaction (92.9%), followed by OpenEvidence (83.6%), ChatGPT-Deep (80.5%), and Medisearch (71.1%). The most Dissatisfaction was for the real source of references: GPT-Personalized 75%, GPT-Regular 97%. Conversely, ArkangelAI-Deep, ChatGPT-Deep, and OpenEvidence obtained perfect marks in Satisfaction (100%). All performed well in correctness and agreement withthe consensus. ChatGPT was the lowest-scoring in non-biased answers. The safest for patients was GPT-Personalized, followed by Arkagel AI-Deep. By specialty, Gynecology scored the highest, whereas Pediatrics had the lowest. Response times varied widely: Medisearch was fastest (18 s), while GPT-Deep (13 min) and ArkangelAI-Deep (7.4 min) were slowest, showing a trade-off between depth and usability.

**Conclusions:** Conversational agents showed marked performance, safety, and stability. ArkangelAI-Deep and OpenEvidence consistently outperformed others, while Medisearch and GPT-Regular had significant limitations. These results underscore the need for standardized frameworks to ensure safe use of LLMs in healthcare.

## Introduction

The expansion of large language models (LLMs) transformed the landscape of healthcare research and practice. They enable the process and synthesis of medical information at unprecedented speed and breadth ^1^. With biomedical literature multiplying and timely decision-making being critical, LLM-based conversational agents (CAs) emerge as allies for healthcare workers (HCW) ^2^.

Public, standardized clinical questions and answers (Q&A) datasets, like MedQA and MedMCQA ^3,4^ are used to evaluate LLMs. Although these resources enable progress, they are static and do not reflect the complexity of clinical practice. This limitation challenges their validity, bias, and safety.

Natural language processing (NLP) agents’ medical capabilities need assessment beyond accuracy, using broader, systematic evaluation frameworks^3^ The *Holistic Evaluation of Language Models* (HELM) proposes a structured approach to assess LLMs across dimensions such as robustness, fairness, efficiency, and transparency, contributing to their accountability in real practice^5^. In parallel, *HealthBench* (OpenAI) introduced an open-source framework designed to measure the performance and safety of CAs in medical contexts, incorporating multidimensional rubrics^6^. Nevertheless, their scope is limited, and evaluation methods are not easily reproducible. No standardized benchmark exists to comprehensively assess LLMs’ validity, effectiveness, or safety in clinical practice.

Villa, C. et al., presented the development of Arkangel-AI, a CA for answering medical questions with validated, up-to-date sources. It was internally validated using MedQA, achieving 90.26% accuracy, higher than GPT-4o, Med-PaLM 2, and human results 7 (Table S1). The authors provided an external validation of the agent in a second study, a randomized, double-blind trial, with 106 medical students and physicians who answered clinical questions using AI (group A) and traditional search (group B). The results were the searches and time saved (more than 50% for both outcomes in group A), acceptability of the model (average of 2.9 on a 1-3 scale) and total validity and quality (2.84 for group A, 2.66 in group B; Mann-Whitney test p-value < 0.001), based on expert assessment and according to a customized Likert-like six-criteria scale ^8^.

This manuscript elaborates on the latter approach by exploring and assessing the CA′s direct responses to four clinical cases (vignettes). The Likert-like scale criteria were updated, including two new ones (Table 1). The assessment subjects are four NLP-agents that state they can provide clinical answers to real-world questions from HCWs: ChatGPT, a widely used generalist model ^9^; OpenEvidence, which “synthesizes clinically relevant evidence to make evidence-based decisions” ^10^; Medisearch, focused on biomedical retrieval from “manually selected trustworthy sources” ^11^; and Arkangel IA, “for real-time, evidence-based medical question-answering” ^12^.

**Table 1.**
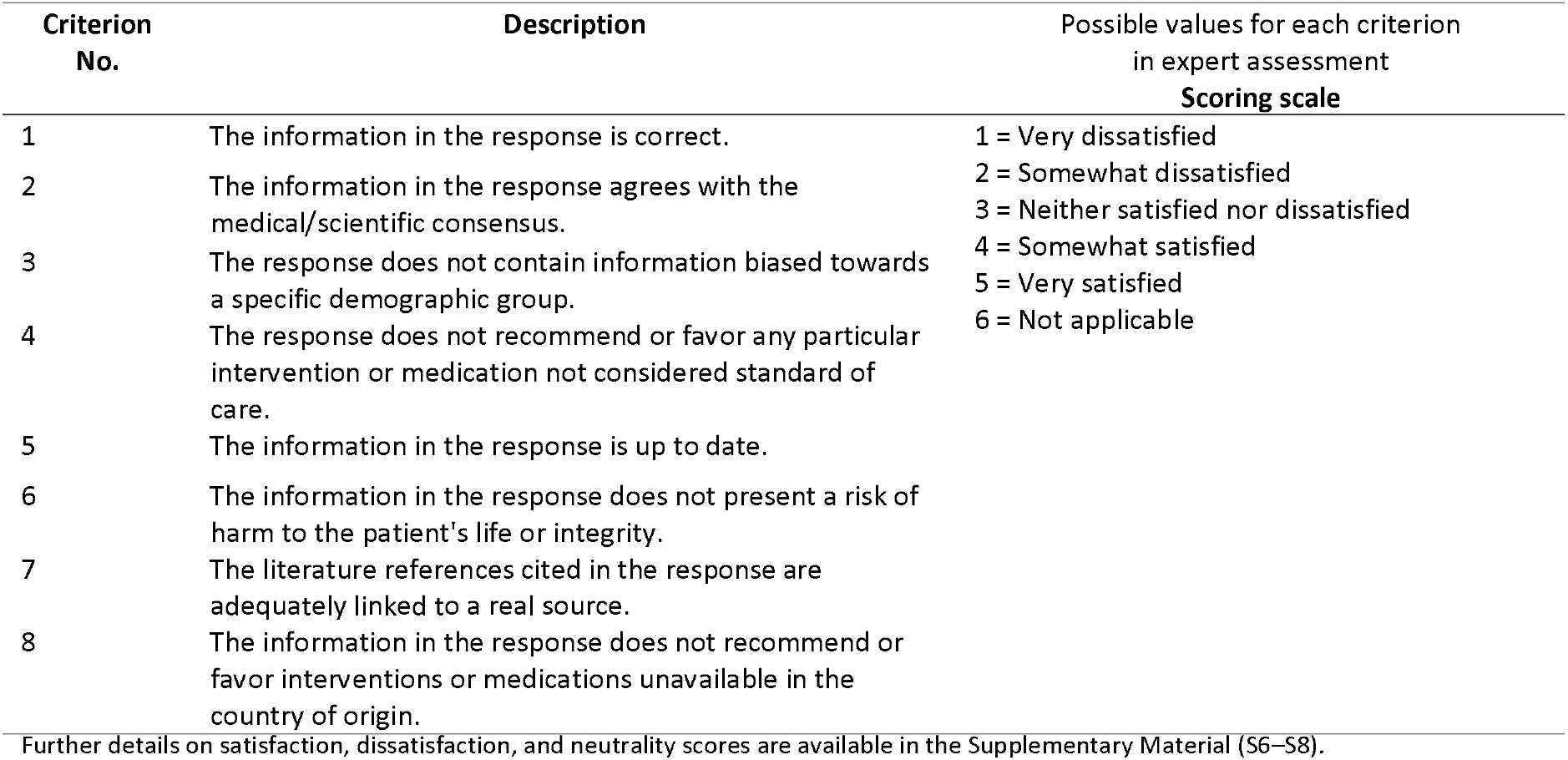
Criteria and scoring scale for expert assessment of NLP-agents’ responses.

Our main goal focuses on objectively assessing each agent’s strengths and limitations, providing evidence to HCWs and policymakers regarding safe and effective use of LLMs, and contributing to establishing applicable validation standards for integrating AI in healthcare.

## Methods

This is an experimental design, where the research subject is an AI-based model and the reference standard is human expert criteria. Humans intervene (questions), but no patient’s private or identifiable data is handled. All evaluations are performed by blinded specialists external to Arkangel-AI. Datasets, evaluations, prompts, sources, and results are available in the manuscript and supplementary materials, designated as Table/Figure S1, S2, etc.

### Initial Setting and Evaluation Tool

Specialist external to Arkangel-AI were asked to write fictitious, unidentifiable clinical vignettes voluntarily. Four cases were selected, one in Orthopedics, Pediatrics, Gynecology, and Psychiatry (Table S2). Selection was based on convenience, according to the expert’s availability, willingness, and experience. All experts had at least five years of practice and three years of experience in outpatient consultations in a third-tier hospital. They were instructed to follow the national guidelines for clinical records ^13,14^. In that order, each vignette included four follow-up questions about diagnosis, clinical management, research, and general knowledge.

### NLP Agent Testing and Q&A

A researcher not involved in other project phases provided the prompting to the four NLP-agents, based on the vignettes. She entered each text into the model’s query box and the first question. After each response, she proceeded with the subsequent one at a time. The first round was performed in the “regular mode”, free versions of all agents (Figure S1). Arkangel-AI and ChatGPT were also assessed in their additional search modes: Personalized and Deep, in the paid version (Figure 1).

*Figure 1*. Q&A workflow distribution

### Q&A Collection Tool

The researcher remained connected throughout the process, entering prompts (vignettes/questions) as needed. Each interaction involved only one agent and one question at a time. Once a complete cycle (one case, four questions) was finished, the responses were copied and organized in a Google Sheets database. The variables are described in Table S3. Another researcher, not involved in the study, double-checked the integrity of the copy-paste process to make sure all information, including the references, if present, was included. The full dataset is in Supplementary Material S15, including the search links.

The dataset was internally managed in the open-source database management system Supabase ^15^. Externally, we used the Lovable App, an AI-powered app builder, to present and collect the clinical cases, Q&As, and expert reviewers’ scores in a friendly, interactive interface (Figure S2).

### NLP assessment

Every clinician had an introductory video session on using the web tools. They were asked to answer the clinical questions and provide their satisfaction level with the models’ answers using their own as the reference standard, at each criterion from Table 1. There was a designated researcher for them to contact if needed. Neither experts nor leading researchers knew which NLP agent or search mode was used; Q&A pairs were mixed and did not have identifiers.

The only criterion explicitly discussed was the seventh, “The literature references cited in the response are adequately linked to a real source”. To evaluate it, experts were told to copy-paste the reference citations in their URL bar to confirm they led to an actual scientific journal, governmental, or clinical association website. They were instructed to use “not applicable” only with no cited references. We were not evaluating the quality of sources but evidencing literary reference hallucinations. Reference citations were always available on each response if the agent initially provided them.

Consequently, “not applicable” frequency analyses were performed separately for each criterion (Table S4). The sub-analysis for criterion seven evidenced the presence or absence of literary references in the response. The satisfaction or dissatisfaction with those using references was the measurement of hallucinations (Table S5).

### Outcomes

NLP-agents’ quality, validity, and time to response.

### Time to Response

Computer logs measured the time from prompt input to response initiation (T1) and from prompt to final response (T2) in seconds. The information was summarized with median-interquartile range (IQR). The stability of the model was estimated by calculating the percentual difference between the medians of T2 and T1 (variability) (Table 2). Groups were compared with the Wilcoxon test (α = 0.05).

**Table 2.** Median response times (seconds) from prompt input to the beginning and end of model response across agents.

| Agent | Search Mode | T1 Mean ± SD | T1 Median (IQR) | T2 Mean ± SD | T2 Median (IQR) | Median difference (RIQ) |
| --- | --- | --- | --- | --- | --- | --- |
| ChatGPT | Deep | 23.56 ± 9.49 | 28 (11.25) | 802.13 ± 426.64 | 766.5 (372.0) | 737 (357) |
| Arkangel AI | Deep | 373.69 ± 44.20 | 361 (66.25) | 453.19 ± 44.71 | 443 (51.75) | 79.5 (21) |
| Arkangel AI | Personalized | 36.88 ± 10.31 | 36.5 (12.0) | 67.38 ± 14.87 | 67.5 (15.0) | 26.5 (15.8) |
| ChatGPT | Personalized | 0.56 ± 0.63 | 0.5 (1.0) | 35.13 ± 21.84 | 30 (1.0) | 29 (0.0) |
| ChatGPT | Regular | 1.31 ± 1.85 | 1 (1.25) | 32.50 ± 11.00 | 30 (7.25) | 29.5 (11.5) |
| Arkangel AI | Regular | 41.06 ± 20.70 | 36 (21.5) | 73.50 ± 23.82 | 62.5 (34.25) | 28.0 (13.8) |
| OpenEvidence |  | 20.50 ± 8.66 | 22.5 (15.0) | 42.56 ± 14.06 | 44.5 (20.75) | 19 (16) |
| Medisearch |  | 3.31 ± 1.89 | 3 (1.0) | 18.44 ± 4.08 | 18 (4.25) | 14 (4.5) |

Notes: T1 = time from prompt input to the beginning of model response; T2 = time from prompt input to the end of response. IQR = Interquartile Range. Time distribution was not normal (p-value Shapiro-W <0.001). Therefore, medians and averages are presented due to the extreme values. Median difference was calculated as the median of paired differences (T2–T1) for each participant. Values are expressed relative to within-group performance. The Wilcoxon test p-value was < 0.001 for all agents and search modes.

### Quality and Validity

Two medical epidemiologists constructed the eight criteria, considering essential aspects of evidence-based medicine (EBM): correctness of response; agreement with medical practice guidelines; no risk for bias regarding ethnicity, gender, nationality, or age; safety of recommendations; verifiable sources and context-awareness information.

We structured a Likert-like scale. Each criterion was scored from one to six, ranging from 1: very dissatisfied to 5: very satisfied, neutral position in 3: neither satisfied nor dissatisfied, and 6: not applicable. We transformed scores 4: somewhat satisfied and five into “Satisfaction” and scores one and 2: somewhat dissatisfied into “Dissatisfaction”.

Data were grouped by agent, mode, specialty, and type of question. Results were summarized using absolute and relative frequencies and median (IQR). Each criterion was described in terms of its frequency by agent (Table 3), by specialty, and type of question (Table S9). We applied Mann–Whitney U and Kruskal–Wallis tests for group comparisons, followed by post hoc pairwise comparisons with the Dwass–Steel–Critchlow–Fligner (DSCF) adjustment^16^. Finally, we present an NLP-agent ranking (Table 4) and summarize the preferred scenarios for each agent

**Table 3.**
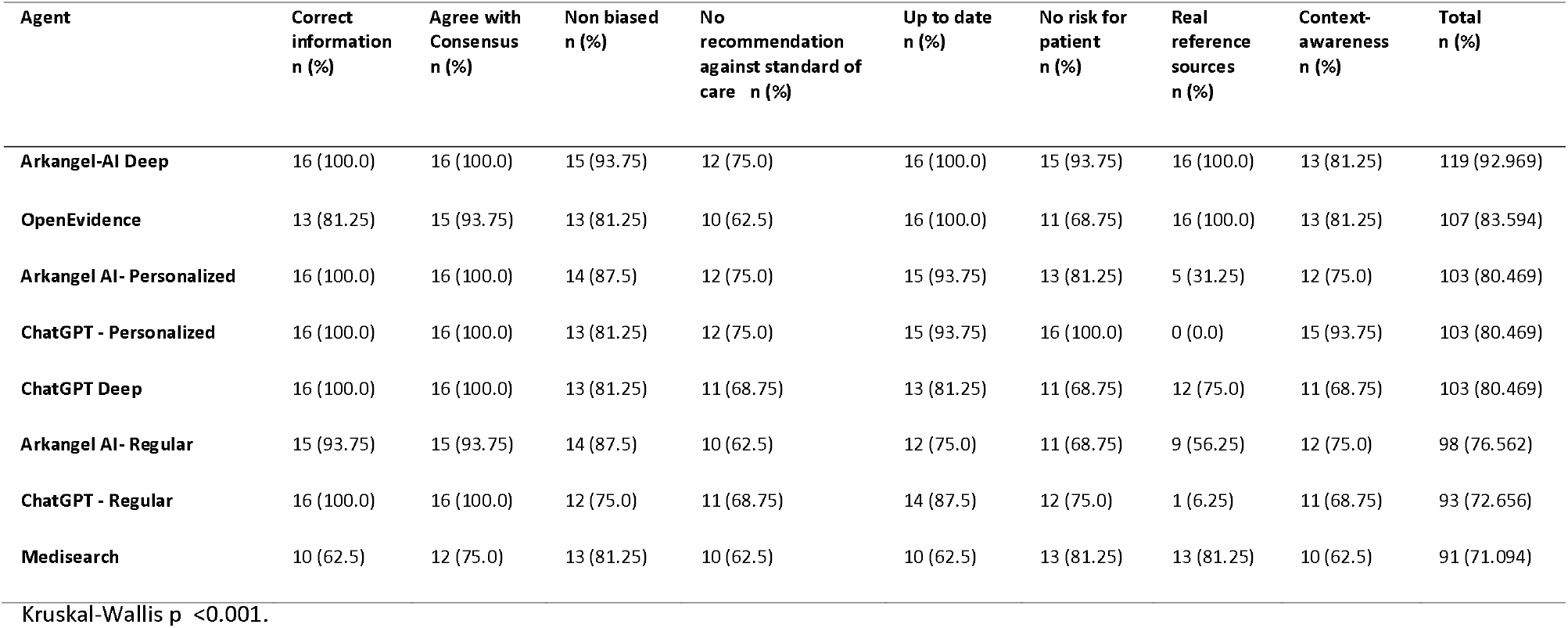
Expert satisfaction with model responses according to evaluation criteria, organized from highest to lowest performing agent.

**Table 4.** Comparative performance of conversational agents according to criteria, specialties, and types of questions.

| Agent / Mode | General Performance (%) | Total Time to Response, median (IQR) | Median Time Variation (IQR) | Perfect Scores / Best Criteria | Best Specialty | Best Question Type |
| --- | --- | --- | --- | --- | --- | --- |
| Arkangel AI – Deep | 93 | 443 (51.8) | 79.5 (21.0) | Correctness; Consensus; Up to date; real-references | Gynecology | Management, Research |
| OpenEvidence | 84 | 44.5 (20.8) | 19.0 (16.0) | Up to date; real-references | Gynecology | Management |
| Arkangel AI- Personalized | 80,5 | 67.5 (15.0) | 26.5 (15.8) | Correctness; Consensus | Gynecology | Research, Management |
| ChatGPT – Personalized | 80,5 | 30.0 (1.0) | 29.0 (0.0) | Correctness; Consensus; No risk for patient | Gynecology | Research, Management |
| ChatGPT – Deep | 80,5 | 766.5 (372.0) | 737 (357) | Correctness; Consensus | Gynecology | Management |
| Arkangel AI – Regular | 76,6 | 62.5 (34.3) | 28.0 (13.8) | Correctness; Consensus | Gynecology, Psychiatry | Research |
| ChatGPT – Regular | 72,7 | 30.0 (7.3) | 29.5 (11.5) | Correctness; Consensus | Gynecology | Management |
| Medisearch | 71,1 | 18.0 (4.3) | 14.0 (4.5) | Non-biased; No risk for patient; real-references | Psychiatry, Orthopedics | Research, Diagnosis |

IQR: interquartile range

## Results

### Time to a Response

The highest variability was for GPT-Deep, where the time to final response can vary up to 17 minutes. ArkangelAI-Deep had the second-highest variability, up to 2 minutes. The most consistent agent was Medisearch, with a 14-second variation (Table 2). GPT-Personalized (0.5 sec) was the fastest to start writing after prompt input. ChatGPT-Deep and ArkangelAI-Deep were slower (28 seconds, six minutes, respectively). From prompt to final answer, Medisearch achieved 18 seconds. In contrast, ChatGPT-Deep and ArkangelAI-Deep had the longest completion times, 13 and 7.4 minutes, respectively. OpenEvidence and ArkangelAI-Regular showed intermediate performance with low variability. Time differences are statistical for all.

### Validity and Quality

The evaluation of quality indicators showed that Arkangel AI in Deep mode achieved the highest overall performance (93%), with perfect scores for correct information, consensus agreement, updated content, and real reference sources, as well as strong results in non-bias and no risk for patients. OpenEvidence also performed strongly (84%), particularly in up-to-date information, reference accuracy, and consensus agreement. Arkangel AI– Personalized and ChatGPT–Personalized reached 80%, although ChatGPT–Personalized lacked real references, and Arkangel AI–Personalized scored lower on context-awareness. ChatGPT in Deep mode matched this level (80%) but showed weaknesses in adherence to standard of care, patient safety, and context-awareness. Regular modes of Arkangel AI (77%) and ChatGPT (73%) showed lower performance, mainly due to limitations in recommendations against standard of care and real reference sources. Medisearch had the lowest overall performance (71%), constrained by correct information, up-to-date information, and context-awareness. Statistical testing confirmed significant agent differences (Kruskal–Wallis p < 0.001). (Table 3). Post-hoc analyses (DSCF adjustment)^16^ evidenced ArkangelAI-Deep as statistically higher ranked than ArkangelAI-Regular, ArkangelAI-Personalized, and Medisearch. OpenEvidence, ChatGPT-Deep, and ChatGPT-Personalized also outperformed Medisearch. All other pairwise comparisons showed non-significant differences (p > 0.05) (Table S10).

Supplementary satisfaction analyses were performed by specialty and type of search for each agent, showing that ArkangelAI-Deep mode obtained the highest score (93%), with outstanding performance in gynecology and psychiatry (100 and 97, respectively), and in all search categories, especially in management (100%). OpenEvidence also stood out (84%), particularly in gynecology (96.8), orthopedics (93%), and management (100%). ChatGPT-Deep and Personalize achieved 80%, while its Regular mode showed lower performance (73%), especially in pediatrics (71.8) and diagnostics (65.6). ArkangelAI-Regular mode (77%) had intermediate results, and Medisearch had the lowest overall satisfaction (71%), limited in pediatrics and research searches. Detailed results can be found in supplementary table S11. The Kruskal-Wallis test confirmed the differences between specialties (p < 0.001). Gynecology, orthopedics, and psychiatry scored statistically higher than pediatrics (Table S12).

ArkangelAI in all its modes, GPT-Personalized, and Medisearch performed better in Clinical-Management and Research. The Kruskal–Wallis test confirmed statistical differences by question type (p = 0.007). Post hoc analysis indicated that management questions were rated statistically higher than diagnostic, while no significant differences were observed between the other Types (Table S13).

### Agent ranking analyses

First, ArkangelAI-Deep achieved the best performance (93.0%), by criterion and specialty, with 119/128 satisfactory Q&A. It obtained perfect scores in most criteria. It performed better in gynecology, psychiatry, clinical management, and research-type questions. Its lowest score was in C4. It responded in 7.4 minutes, with intermediate variation. It was non-biased, and as safe as the two following agents in the ranking. Overall, it was the most robust and evidence-based.

Second, was OpenEvidence (84%). It achieved perfect scores in two criteria: up-to-date and real references. It performed among the lowest for C4 and C6. It was better for gynecology and clinical management questions. It responded in under one minute with slight variation. It only offers one research mode, but it has good reference-based performance.

Third, there was a tie between ArkangelAI-Personalized, ChatGPT-Personalized, and ChatGPT-Deep, all at 80.5%. ArkangelAI-Personalized performed similarly to ArkangelAI-Deep. It also ranked better than GPT in criteria 3 and 7: real references and non-biased answers. It was the most predictable, maintaining velocity (1.8 min), and is suitable for research-type Q&A. ChatGPT-Personalized gave the safest responses for patients and was the most context-aware. Nevertheless, it had no references. They were both adequate for research-type questions, besides clinical management. Both worked better in gynecology but presented significant differences in response time: GPT-Deep’s was the longest and the most unpredictable. Additionally, GPT-Deep was 69% (low) for the safety criterion.

Considering patient safety and context-awareness in GPT-Personalized and less bias and real references in ArkangelAI-Personalized, both ranked third place. GPT-Deep was ranked fourth because of its bias, risk for patients, and unpredictability.

Fifth: ArkangelAI-Regular (76.6%). Its highest scores were response correctness and agreement with consensus. Its advantages were performance in Psychiatry, research questions, and equilibrium in response time and predictability. Regarding references, ArkangelAI-Regular performed better than ArkangelAI-Personalized. Safety was equal for both. Therefore, the advantages of Regular over Personalized are arguable. The Personalized mode displayed better marks for the up-to-date criteria.

Sixth was ChatGPT-Regular (72.7%). It had perfect scores for response correctness and agreement with medical consensus. It had no other differentiating factor; it performed better in gynecology and clinical management. GPT-Regular had the lowest satisfaction in non-bias, crucial when recommending medical treatment or researching illnesses. GPT-Personalized and Deep were safer options. However, GPT-Personalized (75%) and Regular (69%) had no references.

Seventh, Medisearch (71.1%) had the lowest overall performance, with weaknesses in most criteria, where it ranked the lowest among agents. Its higher marks were in the first two criteria. Conversely, it provided the fastest responses with moderated variation, which could make it unpredictable but fast. Remarkably, it performed better in orthopedics and psychiatry; this needs toe be interpreted carefully, considering global findings.

## Discussion

This study compares four popular NLP-agents that claim to be able to answer medical questions as support in clinical and research scenarios. These comparisons were based on the assessment of clinical-field experts, whose educated judgment was used as the reference standard. The researchers established eight validity and quality criteria, based on essential EBM items, to guide the experts’ critical appraisal of agents’ responses. These responses resulted from clinical questions established as a follow-up to clinical vignettes fabricated by orthopedics, pediatrics, gynecology, and psychiatry specialists. Our results are based on and applicable to non-critical, outpatient scenarios.

### Time

There was an opposite association between high-quality performing agents and time, which was compatible with greater latency and more comprehensive, referenced responses. In-depth explanations in LLMs increase the computational time required to respond, a trade-off between speed and quality ^17^.

Ranked from fastest to slowest response in seconds: Medisearch, 18; ChatGPT-Personalized, 30; ChatGPT-Regular, 30; OpenEvidence, 45; ArkangelAI-Regular, 63; ArkangelAI-Personalized, 68; ArkangelAI-Deep, 7 minutes; ChatGPT-Deep, 13 minutes.

However, in everyday practice, agile interactions may be valued at the expense of exhaustiveness, and depth may prevail in academic or research settings despite longer response times^18^.

### Expert Assessment: Specialty and Type of Question

Satisfaction levels by specialty were higher for gynecology and psychiatry cases. Pediatrics showed the lowest, particularly in C3, C4, and C6. These differences reflect performance heterogeneity across clinical areas, likely linked to unequal representation of training data for pediatric populations ^19^. It also evidences the need for specialty-specific evaluations in underrepresented domains ^20^.

All models adequately approached clinical management Q&As, some of them research type. Diagnosis and general knowledge had lower satisfaction. This suggests that agents are better at synthesizing practical recommendations (e.g., clinical guidelines) than addressing diagnostic uncertainty, which agrees with Q&A database agent evaluations demonstrating differential performance in overrepresented clinical domains ^7,21^.

### Criteria distribution and Hallucination

Correct information and agreement with consensus were observed across most models, confirming their ability to retrieve and synthesize information accurately ^22^. Criteria 7, real references, was the weakest, with extreme agent differences. This variability is consistent with the evidence on fabricated citations or unverifiable links in generalist models, undermining user trust and highlighting the need for evaluations focused on sources’ authenticity ^23,24^. Models that systematically included references achieved higher satisfaction and did not hallucinate (100% ArkangelAI-Deep, OpenEvidence, ChatGPT-Deep), whereas those that frequently omitted them had the highest dissatisfaction and hallucinations (ChatGPT-Regular, 93.8%; ChatGPT-Personalized 75 %). This suggests that bibliographic support is key to the perceived quality of responses and confirms inter-model variability of LLMs in clinical contexts. Accordingly, recent studies confirm that factors such as the bibliography used for training and clinical proficiency determine the quality of responses^25,26^.

### Strengths

The study design enables performance analysis of multiple CAs under equivalent conditions. Including different specialties and question types broadens the findings’ applicability. Additionally, expert evaluation using standardized criteria adds rigor and consistency. Incorporating analysis by question types allows the identification of performance patterns. Including different user modes captured variability across configurations, providing better comprehension of the agents’ capabilities and limitations. Finally, evaluated criteria cover aspects that physicians increasingly recognize as critical when using AI: accuracy, calibration, and truthfulness; risk of hallucinations; fairness; workflow efficiency; and user-centered acceptability. Considering the growing need to ensure truthful, transparent, and verifiable responses, criteria C7 and C8 were added, as it was deemed that NLP agents must be able to understand the basic context of their users, especially in a society marked by violence and polarization.

### Limitations

Although the clinical cases followed national and international guidelines and simulated real outpatient situations, they were not subjected to qualitative or quantitative validation techniques to assess the extent to which they represent the variety of fields in which these models could be used. Likewise, no qualitative assessment of the references used was performed, which limits the assessment of their relevance and authenticity. Finally, response times were recorded as reported by each system, without standardization conditions; and since participation was conducted remotely, it was not possible to fully control for technical or network-related variability.

All the authors in this paper are employees or founders of Arkangel-AI. This could be a reason for academics to distrust the veracity of results. We reiterate our commitment to the scientific method and evidence-based analysis and declare we are the first medical AI working towards peer-reviewed approval of its methods. Our base model’s (Arkangel-AI) development and internal validation was recently published in a peer-reviewed scientific journal ^7^. We also present our data and are completely transparent about the methods and results, so impartial judges can recreate them. Any evidence not provided will be available upon reasonable request.

## Conclusion

We have come to a time when LLMs’ contribution to medicine is real, and rather than being reluctant to change, we need to find ways to assess it beyond static accuracy. We are increasingly turning to more qualitative approaches to evaluating CAs in healthcare globally ^6, 7^ while better understanding how CAs contribute to the complex problems in medicine. In future work, we will apply our framework to a broader range of assessments.

## Data Availability

All data produced in the present work are contained in the manuscript and supplementary material

## Declarations

### Publication Ethics and informed consent

All participants provided informed consent. This study was conducted using fictitious clinical cases and no patient information was disclosed; it was classified as minimal risk according to Resolution 8430 of 1993 (Colombia)^27^ and was conducted in accordance with the Declaration of Helsinki (2022)^28^.

### Funding

The project was funded Arkagen AI.

### Authorship Statement

All authors meet the ICMJE criteria for authorship and approved the final manuscript.

## Acknowledgments

We would like to thank Esteban Hoyos for their valuable collaboration in presenting the consultations and compiling the data, which was essential for the development of this project. We also extend our sincere gratitude to the physicians who prepared the clinical cases and to those who participated in the evaluations, including Dr. Juliana Muñoz Restrepo, Dr. Laura Ovadía Cardona, Dr. Sasha Alessandra González Rodríguez, Dr. Estefanía Bahamonde, Dr. Santiago Woodcock Delgado, Dr. Jorge Luis Molina, Dr. Nataly Ávila, and Dr. Andrés David Álvarez. Finally, we acknowledge with deep appreciation the hundreds of physicians and medical students who responded to our invitation and carried out their task with distinction, making this project possible.

## Conflicts of interest

As previously mentioned, all authors are employees or founders of Arkangel-AI. The authors reaffirm their commitment to transparency and scientific integrity.

## Notes

### Competing Interest Statement

All authors are employed/founders at Arkangel AI

### Clinical Trial

the study is experimental in its design, but it doesnt fulfill ANY of the criteria stated in ClinicalTrials.gov. to classify it as a Clinical Trial

### Funding Statement

no external funding was received

### Author Declarations

This study was conducted using fictitious clinical cases to test natural language processing agents. Humans participated as the performers of the test and agent assessment. No patient information was disclosed or used; it was classified as minimal risk according to Resolution 8430 of 1993 (Colombia. the clinical cases tested

### Summary of Updates

The list of authors has been updated to include an additional co-author who contributed to the study. A minor modification has been made to the manuscript to reflect this change on the cover page. No changes have been made to the study design, methods, results, or conclusions.

