## Supplementary material for "Arkangel AI, OpenEvidence, ChatGPT, Medisearch: are they objectively up to medical standards? A real-life assessment of LLMs in healthcare" (from another version of this preprint: version 1, 2025-09-25)

**Supplementary Materials**

**Supplementary Table S1.** Benchmark comparisons.

Reported SoTA LLMs on MedQA (USMLE) test set, including Arkangel AI.

| Model | Test set questions used (n) | Accuracy |
|---|---|---|
| **Arkangel AI v3** | 1273 | 90.26% |
| **GPT-4o** | 1273 | 87.51% |
| **GPT-4o mini** | 1273 | 73.61% |
| **Med-PaLM 2** | 1273 | 85.40% |
| **GPT-4 (Medprompt)** | 255 | 90.20% |
| **Human experts** | 1273 | 90.20% |

**Supplementary Table S2.** Clinical vignettes and derived questions.

| Speciality | Clinical case |
|---|---|
| **Gynecology** | A woman in her early thirties, married and nulliparous, currently enrolled in a technical program and employed in a service-related job, seeks urgent evaluation for a one-month history of low mood, initial insomnia, loss of energy, and frequent nightmares ("I have nightmares almost every night"). She denies loss of appetite. She reports distractibility and long-standing passive thoughts of death beginning in adolescence. She had past episodes of suicidal ideation, the last one several years ago. She acknowledges emotional instability. In the past, she responded to frustration with self-injurious and aggressive behaviors ("I burned myself with cigarettes, punched walls, and tried to divert attention to something physical"). She also reports decreased libido.<br><br>Over the last two years, she has increased her alcohol consumption with loss of control ("we could drink for 2–3 days straight"), leading to functional impairment ("I've put myself at risk because I have blackouts") and relationship difficulties. She also uses marijuana and tobacco regularly. She has been undergoing marital separation in the past month. She reports previous episodes with similar characteristics. She describes alternating affective episodes of increased energy and productivity followed by periods of slowing and low motivation. For the past two months she has also felt restless and anxious. She denies current or past hallucinations, delusional ideas, or persecutory thoughts.<br><br>Past history: Respiratory allergies. No surgeries. No known drug allergies.<br>Substance use: Cigarette use since adolescence, currently daily; vaping in recent years with limited benefit. Cannabis since late adolescence, now several times per week. Remote experimental use of cocaine, hallucinogens, and other substances; no recent use.<br>Family history: Relatives with depression, suicide, and substance use disorders. |
| **Question type** | |

**Diagnosis** What is the most likely diagnosis in this patient?

**Management** What is the appropriate management for this patient according to current guidelines from the National Institute of Health (Instituto Nacional de Salud, INS)?

**Research** What is the efficacy of HPV vaccination in preventing cervical cancer among women who did not receive the vaccine during adolescence?

**General knowledge** What role does a positive family history play in this condition?

**Orthopedics** Personal data: Female, adult in her early thirties, single, with incomplete formal education, currently a homemaker.

Chief complaint: "Persistent neck pain."

History of present illness: The patient reports a history of cervical pain of approximately two months, with mechanical features, particularly noticeable during neck rotation and extension. The pain is intermittent and associated with a sensation of head instability and occasional dizziness, described as feeling as if her head were "floating" or "detached" from her body. Symptoms worsen with routine activities such as reading or watching television and are also exacerbated by rapid side-to-side neck movements. Recently, she has experienced pain radiating to the shoulders and occasional numbness in the fingers. Her medical background includes autoimmune disease and chronic musculoskeletal disorders that could contribute to these symptoms. The neck pain has increasingly limited daily activities, and usual analgesics have become less effective.

Past medical history: Autoimmune disease with inflammatory arthritis of large and small joints; irregular treatment with corticosteroids and disease-modifying agents. Metabolic disorder requiring insulin therapy, with irregular adherence. Chronic anemia with occasional transfusions. Chronic arthritis in wrists, knees, elbows, and ankles; mechanical hip pain with restricted mobility.

Physical examination: The patient is in acceptable general condition, alert and oriented. Musculoskeletal exam: Limited cervical spine rotation; swelling and warmth in some joints (elbows, metacarpophalangeal joints); muscle atrophy of the interossei in both hands. Spurling test: Negative, without clear radiculopathy. Cervical palpation: Localized tenderness with reduced range of head motion. Neurologic exam: No major motor or sensory deficits, though the patient describes mild paresthesia in the fingers.

Diagnostic studies: Cervical radiograph: Evidence of mild atlantoaxial (C1–C2) instability, more noticeable during movement. Cervical MRI: Ligamentous and degenerative changes in the atlantoaxial complex, suggesting instability. Electromyography: No evidence of significant cervical radiculopathy.

**Question type**

**Diagnosis** What is the most likely diagnosis from an orthopedic standpoint?

**Management** What are the surgical indications in patients with atlantoaxial instability of the cervical spine?

**Research** What is the efficacy and safety of cervical collars in patients with cervical spine pathology?

**General knowledge** What recommendations and restrictions should be followed for therapeutic exercise and physical therapy in a patient with cervical spine pathology?

| | |
|---|---|
| **Pediatrics** | A female infant of approximately two months of age, exclusively breastfed on demand, has a one-month history of frequent post-feed emesis and persistent straining. During the past week, the stools have shown streaks of blood. She has not received formula or complementary foods.<br>The perinatal history corresponds to a full-term newborn delivered by cesarean section without complications, with unremarkable prenatal care and normal obstetric ultrasounds. Birth weight was appropriate for gestational age, and the infant has received the immunizations recommended for her age.<br>Growth assessment shows a weight-for-length of −2 standard deviations, which represents a decline from a previous measurement of −1 SD. Weight-for-age is −1 SD and length-for-age is −0.5 SD, indicating faltering growth with downward crossing of curves, mainly affecting weight.<br><br>On physical examination, vital signs are within normal limits. The infant is well hydrated, alert, and interactive. Mucous membranes are moist, without jaundice, and the head is normocephalic. Chest expansion is symmetric, without retractions, and breath sounds are clear bilaterally. Cardiac auscultation reveals a regular rhythm with no murmurs. The abdomen is soft and non-distended, without palpable masses or organomegaly, and the umbilicus is normal. Palpation does not produce apparent pain. Perianal inspection shows a normal anus, without fissures or skin tags. The skin reveals mild eczema on the malar region and faint eczema in the antecubital fossae. Neurologic evaluation demonstrates normal tone and reflexes for age.<br><br>In summary, this is an exclusively breastfed infant with growth faltering, a one-month history of post-feed emesis and straining, and new-onset hematochezia, accompanied by mild eczema and otherwise unremarkable systemic findings. |
| **Question type** | |
| **Diagnosis** | What is the most likely diagnosis in this patient? |
| **Management** | What feeding options are recommended for the management of infants with this diagnosis? |
| **Research** | What is the effectiveness of maternal cow's-milk elimination diets during exclusive breastfeeding for managing cow's-milk protein allergy in infants? |
| **General knowledge** | How can proteins from the maternal diet trigger an allergic response in exclusively breastfed infants? |
| **Psychiatry** | A woman in her early thirties, married and nulliparous, currently enrolled in a technical program and employed in a service-related job, seeks urgent evaluation for a one-month history of low mood, initial insomnia, loss of energy, and frequent nightmares, which she describes as occurring almost every night. She denies loss of appetite. She reports distractibility and longstanding passive thoughts of death beginning in adolescence. She acknowledges a history of suicidal ideation, the most recent episode occurring several years ago. She describes herself as emotionally unstable and notes that in the past she reacted to frustration with aggression and self-injurious behavior, such as burning herself with cigarettes or punching walls. She also reports decreased libido.<br><br>Over the past two years she has increased her alcohol consumption with loss of control, sometimes drinking for two to three days consecutively. This pattern has resulted in functional impairment, including blackouts that she perceives as putting her at risk, and has contributed to marital conflict. She also reports regular use of marijuana and tobacco. She is currently undergoing marital separation and describes similar episodes in the past. She reports alternating periods of heightened affective activation, with a sense of being "revved up" and highly productive, |

followed by episodes of psychomotor slowing and low energy. For the past two months she has also experienced restlessness and anxiety. She denies current or past hallucinations, delusional ideas, grandiosity, or persecutory thoughts.

Her past medical history is notable for asthma and allergic rhinitis. She has no history of surgeries and no known drug allergies. Regarding substance use, she began smoking cigarettes in adolescence, with heavy use in the past five years; currently she smokes about ten cigarettes per day and has also been vaping in an attempt to reduce consumption, without success. Cannabis use began in late adolescence, with a daily pattern established in early adulthood; she currently consumes several joints per week, with the longest abstinence lasting only ten days, a few months ago. She reports remote experimental use of cocaine, hallucinogens, and LSD, but denies any use of MDMA.

Her family history is significant for mood disorders and substance use: one relative died by suicide, another had depression, and her parents have histories of alcohol and cannabis abuse.

| **Question type** | |
|---|---|
| **Diagnosis** | What are the most likely diagnoses in this patient? |
| **Management** | What pharmacologic and non-pharmacologic management is recommended for this patient? |
| **Research** | What is the effectiveness of integrated treatment programs for bipolar disorder and substance use disorder in reducing relapses and improving functioning in adults? |
| **General knowledge** | How does bipolar disorder differ from major depression and from schizophrenia? |

**Supplementary Figure S1**. Landing pages of NLP-based tools evaluated.

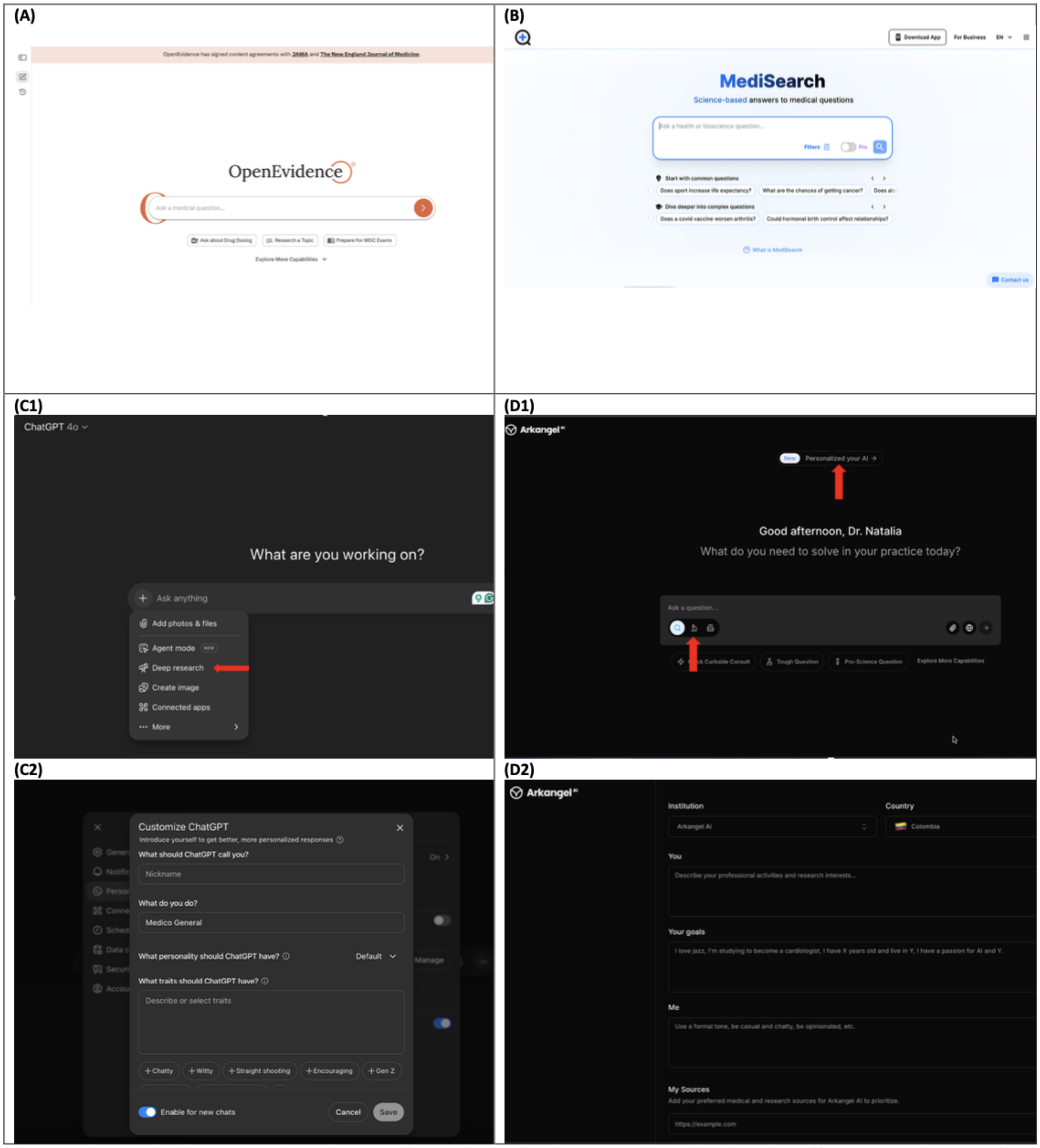

(A).OpenEvidence. Taken from: OpenEvidence homepage [Internet]. 2025 [cited 2025 Sept 8]. Available from: https://www.openevidence.com/
(B).MediSearch. Taken from: MediSearch homepage [Internet]. 2025 [cited 2025 Sept 8]. Available from: https://medisearch.io/
(C). OpenAI. Taken from: ChatGPT—Deep Research; Personalization [Internet]. 2025 [cited 2025 Sept 8]. Available from: https://chatgpt.com/
(D).Arkangel AI. Taken from: Arkangel AI—Deep Reasoning; Personalized—homepage [Internet]. 2025 [cited 2025 Sept 8]. Available from: https://arkangel.ai/

**Supplementary Table S3.** Variables included in the evaluation dataset

| Number | Variables in columns | Description of possible values |
|---|---|---|

| | | |
|---|---|---|
| 1 | NLP Agent | ChatGPT (GPT); OpenEvidence (OE); Medisearch (Ms); Arkangel AI (Arkangel AI) |
| 2 | Search Mode | Regular; Deep; Personalized |
| 3 | Case Specialty | Orthopedics; Pediatrics; Gynecology; Psychiatry |
| 4 | Type of Question | Diagnosis; Management; Research; General knowledge |
| 5 | Agent Response | Text answered by the corresponding agent |
| 6 | Time | Two columns: (a) seconds from prompt input to beginning of model response; (b) seconds to end of response |
| 7 | Expert Satisfaction | Likert-like scale across eight quality criteria (6 scores, see Table 2), assigned by specialists; blinded to agent and mode |

*The original dataset is found in Supplementary Material 15.

**Supplementary Figure S2**. Lovable App personalized landing page used for the evaluation.

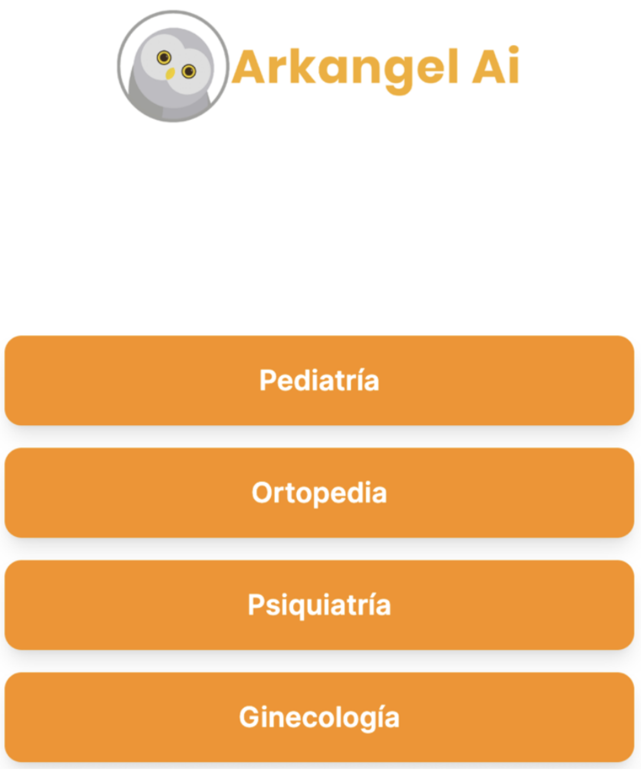

*The study was conducted in Spanish
Take from: https://preview--arkangel-med-forms.lovable.app/

**Supplementary Table S4.** Frequency of “not applicable” responses by evaluation criterion.

| Agent | Search type | Correct information n (%) | Agrees with consensus n (%) | Non biased n (%) | No recommendation against standard of care n (% | Up to date n (%) | No risk for patient n (%) | Real reference sources n (%) | Context-awareness n (%) | Total n (%) |
|---|---|---|---|---|---|---|---|---|---|---|
| **Arkangel AI** | **Regular** | 0 (0) | 0 (0) | 1 (6.2) | 6 (37.5) | 0 (0) | 5 (31.2) | 1 (6.2) | 4 (25.0) | **17 (13.3)** |
| | **Personalized** | 0 (0) | 0 (0) | 2 (12.5) | 4 (25.0) | 0 (0) | 0 (0) | 3 (18.8) | 4 (25.0) | **13 (11.6)** |
| | **Deep** | 0 (0) | 0 (0) | 1 (6.2) | 4 (25.0) | 0 (0) | 1 (6.2) | 0 (0) | 5 (31.2) | **11 (8.6)** |
| **ChatGPT** | **Regular** | 0 (0) | 0 (0) | 4 (25.0) | 5 (31.2) | 0 (0) | 4 (25.0) | 11 (68.8) | 5 (31.2) | **29 (22.7)** |
| | **Personalized** | 0 (0) | 0 (0) | 3 (18.8) | 4 (25.0) | 0 (0) | 0 (0) | 12 (75.0) | 1 (6.2) | **20 (15.6)** |
| | **Deep** | 0 (0) | 0 (0) | 3 (18.8) | 5 (31.2) | 0 (0) | 0 (0) | 0 (0) | 5 (31.2) | **13 (10.2)** |
| **OpenEvidence** | **Regular** | 0 (0) | 0 (0) | 2 (12.5) | 6 (37.5) | 0 (0) | 5 (31.2) | 0 (0) | 3 (18.8) | **16 (12.5)** |
| **Medisearch** | **Regular** | 0 (0) | 0 (0) | 2 (12.5) | 5 (31.2) | 0 (0) | 2 (12.5) | 0 (0) | 6 (37.5) | **15 (11.7)** |

**Supplementary Table S5.** Presence of reference citations and associated satisfaction across agents and modes.

| Agent | Search type | Present reference | | Reference not present n=27 (%) |
|---|---|---|---|---|
| | | Satisfied n=80 (%) | Not satisfied n=48 (%) | |
| **Arkangel AI** | **Regular** | 9 (56.25) | 7 (43.75) | 1 (6.25) |
| **Arkangel AI** | **Personalized** | 5 (31.25) | 11 (68.75) | 3 (18.75) |
| **Arkangel AI** | **Deep** | 16 (100.0) | 0 (0.0) | 0 (0.0) |
| **ChatGPT** | **Regular** | 1 (6.25) | 15 (93.75) | 11 (68.75) |
| **ChatGPT** | **Personalized** | 4 (25.0) | 12 (75.0) | 12 (75.0) |
| **ChatGPT** | **Deep** | 16 (100.0) | 0 (0.0) | 0 (0.0) |
| **OpenEvidence** | **Regular** | 16 (100.0) | 0 (0.0) | 0 (0.0) |
| **Medisearch** | **Regular** | 13 (81.25) | 3 (18.75) | 0 (0.0) |

**Supplementary Table S6.** Disaggregated satisfaction scores by criterion and agent

| Agent | Search type | Correct information n (%) | | | Agrees with consensus n (%) | | | Non biased n (%) | | | No recommendation against standard of care n (%) | | | Up to date n (%) | | | No risk for patient n (%) | | | Real reference sources n (%) | | | Context-awareness n (%) | | | Total n | Total Satisfaction |
|---|---|---|---|---|---|---|---|---|---|---|---|---|---|---|---|---|---|---|---|---|---|---|---|---|---|---|---|
| | | Satisfaction n=118 (92.188) | | | Satisfaction n=112 (95.312) | | | Satisfaction n=107 (83.594) | | | Satisfaction n=88 (68.75) | | | Satisfaction n=111 (86.719) | | | Satisfaction n=102 (79.688) | | | Satisfaction n=72 (56.25) | | | Satisfaction n=97 (75.781) | | | | |
| | | Somewhat satisfied n (%) | Very satisfied n (%) | Median (RIQ) | Somewhat satisfied n (%) | Very satisfied n (%) | Median (RIQ) | Somewhat satisfied n (%) | Very satisfied n (%) | Median (RIQ) | Somewhat satisfied n (%) | Very satisfied n (%) | Median (RIQ) | Somewhat satisfied n (%) | Very satisfied n (%) | Median (RIQ) | Somewhat satisfied n (%) | Very satisfied n (%) | Median (RIQ) | Somewhat satisfied n (%) | Very satisfied n (%) | Median (RIQ) | Somewhat satisfied n (%) | Very satisfied n (%) | Median (RIQ) | | |
| **Arkangel AI** | **Regular** | 9 (60.0) | 6 (40.0) | 4 (1) | 6 (40.0) | 9 (60.0) | 5 (1) | 4 (28.6) | 10 (71.4) | 5 (1) | 2 (20.0) | 8 (80.0) | 5 (0) | 3 (25.0) | 9 (75.0) | 5 (1) | 2 (18.2) | 9 (81.8) | 5 (0) | 5 (55.6) | 4 (44.4) | 4 (1) | 1 (8.3) | 11 (91.7) | 5 (0) | 98 | 76,56 |
| | **Personalized** | 7 (43.8) | 9 (56.2) | 5 (1) | 6 (37.5) | 10 (62.5) | 5 (1) | 5 (35.7) | 9 (64.3) | 5 (0) | 1 (8.3) | 11 (91.7) | 5 (0) | 9 (60.0) | 6 (40.0) | 4 (1) | 1 (7.7) | 12 (92.3) | 5 (0) | 1 (20.0) | 4 (80.0) | 5 (0) | 1 (8.3) | 11 (91.7) | 5 (0) | 103 | 80,47 |
| | **Deep** | 3 (18.8) | 13 (81.2) | 5 (0) | 0 (0.0) | 16 (100.0) | 5 (0) | 1 (6.7) | 14 (93.3) | 5 (0) | 0 (0.0) | 12 (100.0) | 5 (0) | 1 (6.2) | 15 (93.8) | 5 (0) | 0 (0.0) | 15 (100.0) | 5 (0) | 2 (12.5) | 14 (87.5) | 5 (0) | 0 (0.0) | 13 (100.0) | 5 (0) | 119 | 92,97 |
| **ChatGPT** | **Regular** | 5 (31.2) | 11 (68.8) | 5 (1) | 4 (25.0) | 12 (75.0) | 5 (0) | 0 (0.0) | 12 (100.0) | 5 (0) | 1 (9.1) | 10 (90.9) | 5 (0) | 5 (35.7) | 9 (64.3) | 5 (0) | 2 (16.7) | 10 (83.3) | 5 (0) | 1 (100.0) | 0 (0.0) | 4 (0) | 1 (9.1) | 10 (90.9) | 5 (0) | 98 | 72,66 |
| | **Personalized** | 1 (6.2) | 15 (93.8) | 5 (0) | 1 (6.2) | 15 (93.8) | 5 (0) | 1 (7.7) | 12 (92.3) | 5 (0) | 0 (0.0) | 12 (100.0) | 5 (0) | 3 (20.0) | 12 (80.0) | 5 (0) | 0 (0.0) | 16 (100.0) | 5 (0) | 0 (0.0) | 0 (0.0) | 0 (0) | 1 (6.7) | 14 (93.3) | 5 (0) | 103 | 80,47 |
| | **Deep** | 2 (12.5) | 14 (87.5) | 5 (0) | 2 (12.5) | 14 (87.5) | 5 (0) | 2 (15.4) | 11 (84.6) | 5 (0) | 0 (0.0) | 11 (100.0) | 5 (0) | 4 (30.8) | 9 (69.2) | 5 (0) | 0 (0.0) | 11 (100.0) | 5 (0) | 8 (50.0) | 4 (25.0) | 4 (1) | 1 (9.1) | 10 (90.9) | 5 (0) | 103 | 80,47 |
| **OpenEvidence Regular** | | 4 (30.8) | 9 (69.2) | 5 (1) | 1 (6.7) | 14 (93.3) | 5 (0) | 0 (0.0) | 13 (100.0) | 5 (0) | 0 (0.0) | 10 (100.0) | 5 (0) | 4 (25.0) | 12 (75.0) | 5 (0) | 1 (9.1) | 10 (90.9) | 5 (0) | 2 (12.5) | 14 (87.5) | 5 (0) | 0 (0.0) | 13 (100.0) | 5 (0) | 107 | 83,59 |
| **Medisearch Regular** | | 9 (90.0) | 1 (10.0) | 4 (0) | 8 (66.7) | 4 (33.3) | 4 (1) | 5 (38.5) | 8 (61.5) | 5 (0) | 4 (40.0) | 6 (60.0) | 5 (0) | 3 (30.0) | 7 (70.0) | 5 (0) | 5 (38.5) | 8 (61.5) | 5 (0) | 4 (30.8) | 9 (69.2) | 5 (0) | 2 (20.0) | 8 (80.0) | 5 (0) | 91 | 71,09 |

**Supplementary Table S7**. Disaggregated dissatisfaction scores by criterion and agent

| Agent | Search type | Correct information n (%) | | | Agrees with consensus n (%) | | | Non biased n (%) | | | No recommendation against standard of care n (%) | | | Up to date n (%) | | | No risk for patient n (%) | | | Real reference sources n (%) | | | Context-awareness n (%) | | | |
|---|---|---|---|---|---|---|---|---|---|---|---|---|---|---|---|---|---|---|---|---|---|---|---|---|---|---|
| | | Dissatisfied | | | Dissatisfied | | | Dissatisfied | | | Dissatisfied | | | Dissatisfied | | | Dissatisfied | | | Dissatisfied | | | Dissatisfied | | | |
| | | Somewhat dissatisfied n (%) | Very dissatisfied n (%) | Median (RIQ) | Somewhat dissatisfied n (%) | Very dissatisfied n (%) | Median (RIQ) | Somewhat dissatisfied n (%) | Very dissatisfied n (%) | Median (RIQ) | Somewhat dissatisfied n (%) | Very dissatisfied n (%) | Median (RIQ) | Somewhat dissatisfied n (%) | Very dissatisfied n (%) | Median (RIQ) | Somewhat dissatisfied n (%) | Very dissatisfied n (%) | Median (RIQ) | Somewhat dissatisfied n (%) | Very dissatisfied n (%) | Median (RIQ) | Somewhat dissatisfied n (%) | Very dissatisfied n (%) | Median (RIQ) | Total dissatisfied |
| **Arkangel AI** | **Regular** | 1 (6.2) | 0 (0.0%) | 2 (0) | 0 (0.0) | 0 (0.0) | 0 (0) | 0 (0.0) | 0 (0.0) | 0 (0) | 0 (0.0) | 0 (0.0) | 0 (0) | 2 (12.5) | 0 (0.0) | 2.0 (0) | 0 (0.0) | 0 (0.0) | 0 (0) | 3 (18.8) | 1 (6.2) | 2.0 (0) | 0 (0.0) | 0 (0.0) | 0 (0) | 5,47 |
| | **Personalized** | 0 (0.0%) | 0 (0.0%) | 0 (0) | 0 (0.0) | 0 (0.0) | 0 (0) | 0 (0.0) | 0 (0.0) | 0 (0) | 0 (0.0) | 0 (0.0) | 0 (0) | 0 (0.0) | 1 (6.2) | 1 (0) | 0 (0.0) | 0 (0.0) | 0 (0) | 6 (37.5) | 0 (0.0) | 2.0 (0) | 0 (0.0) | 0 (0.0) | 0 (0) | 5,47 |
| | **Deep** | 0 (0.0%) | 0 (0.0%) | 0 (0) | 0 (0.0) | 0 (0.0) | 0 (0) | 0 (0.0) | 0 (0.0) | 0 (0) | 0 (0.0) | 0 (0.0) | 0 (0) | 0 (0.0) | 0 (0.0) | 0 (0) | 0 (0.0) | 0 (0.0) | 0 (0) | 0 (0.0) | 0 (0.0) | 0 (0) | 0 (0.0) | 0 (0.0) | 0 (0) | 0,00 |
| **ChatGPT** | **Regular** | 0 (0.0%) | 0 (0.0%) | 0 (0) | 0 (0.0) | 0 (0.0) | 0 (0) | 0 (0.0) | 0 (0.0) | 0 (0) | 0 (0.0) | 0 (0.0) | 0 (0) | 1 (6.2) | 0 (0.0) | 2 (0) | 0 (0.0) | 0 (0.0) | 0 (0) | 1 (6.2) | 2 (12.5) | 1 (1) | 0 (0.0) | 0 (0.0) | 0 (0) | 3,13 |
| | **Personalized** | 0 (0.0%) | 0 (0.0%) | 0 (0) | 0 (0.0) | 0 (0.0) | 0 (0) | 0 (0.0) | 0 (0.0) | 0 (0) | 0 (0.0) | 0 (0.0) | 0 (0) | 0 (0.0) | 0 (0.0) | 0 (0) | 0 (0.0) | 0 (0.0) | 0 (0) | 0 (0.0) | 2 (12.5) | 1.0 (0) | 0 (0.0) | 0 (0.0) | 0 (0) | 1,56 |
| | **Deep** | 0 (0.0%) | 0 (0.0%) | 0 (0) | 0 (0.0) | 0 (0.0) | 0 (0) | 0 (0.0) | 0 (0.0) | 0 (0) | 0 (0.0) | 0 (0.0) | 0 (0) | 0 (0.0) | 0 (0.0) | 0 (0) | 0 (0.0) | 0 (0.0) | 0 (0) | 1 (6.2) | 0 (0.0) | 2 (0) | 0 (0.0) | 0 (0.0) | 0 (0) | 0,78 |
| **OpenEvidence Regular** | | 4 (30.8) | 2 (12.5%) | 0 (0.0%) | 2 (0) | 1 (6.2) | 0 (0.0) | 2 (0) | 0 (0.0) | 0 (0.0) | 0 (0) | 0 (0.0) | 0 (0.0) | 0 (0) | 0 (0.0) | 0 (0.0) | 0 (0) | 0 (0.0) | 0 (0.0) | 0 (0) | 0 (0.0) | 0 (0.0) | 0 (0) | 0 (0.0) | 0 (0.0) | 2,34 |
| **Medisearch Regular** | | 9 (90.0) | 4 (25.0%) | 1 (6.2%) | 2 (0) | 3 (18.8) | 0 (0.0) | 2 (0) | 1 (6.2) | 0 (0.0) | 2 (0) | 1 (6.2) | 0 (0.0) | 2 (0) | 3 (18.8) | 2 (12.5) | 2 (1) | 0 (0.0) | 0 (0.0) | 0 (0) | 2 (12.5) | 1 (6.2) | 2 (1) | 0 (0.0) | 0 (0.0) | 14,84 |

**Supplementary Table S8**. Neutral responses ("Neither agree nor disagree") by agent and evaluation criterion

| Agent | Search type | Correct information n(%) | Agree with Consensus n (%) | Non biased n(%) | No recommendation against standard of care n (%) | Up to date n (%) | No risk for patient n(%) | Real reference sources n(%) | Context-awareness n (%) | Total n(%) |
|---|---|---|---|---|---|---|---|---|---|---|
| **Arkangel AI** | **Regular** | 0 (0.0) | 1 (0.8) | 1 (0.8) | 0 (0.0) | 2 (1.6) | 1 (0.8) | 2 (1.6) | 0 (0.0) | 7 (0,055) |
| | **Personalized** | 0 (0.0) | 0 (0.0) | 0 (0.0) | 0 (0.0) | 1 (0.8) | 0 (0.0) | 1 (0.8) | 0 (0.0) | 2 (0.016) |
| | **Deep** | 1 (0.8) | 0 (0.0) | 1 (0.8) | 0 (0.0) | 0 (0.0) | 0 (0.0) | 0 (0.0) | 0 (0.0) | 3 (0.016) |
| **ChatGPT** | **Regular** | 0 (0.0) | 0 (0.0) | 0 (0.0) | 0 (0.0) | 0 (0.0) | 0 (0.0) | 0 (0.0) | 0 (0.0) | 0 (0.0) |
| | **Personalized** | 0 (0.0) | 0 (0.0) | 0 (0.0) | 0 (0.0) | 3 (2.3) | 0 (0.0) | 3 (2.3) | 0 (0.0) | 6 (0.047) |
| | **Deep** | 0 (0.0) | 0 (0.0) | 0 (0.0) | 0 (0.0) | 0 (0.0) | 0 (0.0) | 2 (1.6) | 0 (0.0) | 2 (0.016) |
| **OpenEvidence Regular** | | 0 (0.0) | 0 (0.0) | 0 (0.0) | 0 (0.0) | 1 (0.8) | 0 (0.0) | 2 (1.6) | 0 (0.0) | 3 (0.023) |
| **Medisearch Regular** | | 1 (0.8) | 1 (0.8) | 0 (0.0) | 0 (0.0) | 1 (0.8) | 1 (0.8) | 0 (0.0) | 0 (0.0) | 4 (0.031) |

**Supplementary Table S9**. Distribution of Specialists' Satisfaction by Specialty and question type, according to evaluation criterion.

| Specialty | Correct information n(%) | Agrees with consensus n(%) | Non biased n(%) | No recommendation against standard of care n (%) | Up to date n(%) | No risk for patient n(%) | Real reference sources n(%) | Context-awareness n (%) | Total n(%) |
|---|---|---|---|---|---|---|---|---|---|
| **Gynecology** | 29 (90.625) | 31 (96.875) | 31 (96.875) | 32 (100.0) | 25 (78.125) | 32 (100.0) | 21 (65.625) | 32 (100.0) | 233(91,016) |
| **Orthopedics** | 32 (100.0) | 32 (100.0) | 28 (87.5) | 24 (75.0) | 32 (100.0) | 28 (87.5) | 16 (50.0) | 25 (78.125) | 217 (84,766) |

| | | | | | | | | | |
|---|---|---|---|---|---|---|---|---|---|
| **Pediatrics** | 27 (84.375) | 28 (87.5) | 17 (53.125) | 10 (31.25) | 22 (68.75) | 12 (37.5) | 16 (50.0) | 11 (34.375) | 143 (55,859) |
| **Psychiatry** | 30 (93.75) | 31 (96.875) | 31 (96.875) | 22 (68.75) | 32 (100.0) | 30 (93.75) | 19 (59.375) | 29 (90.625) | 224 (87,500) |
| **Question** | | | | | | | | | |
| **Research** | 28 (87.5) | 31 (96.875) | 27 (84.375) | 26 (81.25) | 29 (90.625) | 25 (78.125) | 21 (65.625) | 24 (75.0) | 211 (82.421) |
| **Diagnosis** | 31 (96.875) | 31 (96.875) | 25 (78.125) | 10 (31.25) | 27 (84.375) | 23 (71.875) | 18 (56.25) | 19 (59.375) | 184 (71.875) |
| **Management** | 28 (87.5) | 29 (90.625) | 32 (100.0) | 31 (96.875) | 28 (87.5) | 30 (93.75) | 17 (53.125) | 32 (100.0) | 227 (88.671) |
| **General Knowledge** | 31 (96.875) | 31 (96.875) | 23 (71.875) | 21 (65.625) | 27 (84.375) | 24 (75.0) | 16 (50.0) | 22 (68.75) | 195 (76.171) |

**Supplementary Table S10**. Post hoc significance tests of expert satisfaction between models

| Comparison Dwass steel-Critclow-Figner | **p-value** |
|---|---|
| Arkangel AI Regular vs Arkangel AI Deep | 0.016 |
| OpenEvidence Regular vs Medisearch Regular | 0.003 |
| Arkangel AI Deep vs Arkangel AI Personalized | 0.004 |
| Arkangel AI Deep vs Medisearch Regular | <0.001 |
| ChatGPT Deep vs Medisearch Regular | 0.036 |
| ChatGPT Personalized / Regular vs Medisearch Regular | 0.003 |

Notes: Other pairs >0.05

**Supplementary Table S11.** Distribution of expert satisfaction by specialty, question type, and AI agent

| **Agent** | **Arkangel AI** | | | **ChatGPT** | | | **Medisearch** | **OpenEvidence** |
|---|---|---|---|---|---|---|---|---|
| | **Deep** | **Personalized** | **Regular** | **Deep** | **Personalized** | **Regular** | **Regular** | **Regular** |

| | | | | | | | | |
|---|---|---|---|---|---|---|---|---|
| **Specialty** | | | | | | | | |
| **Gynecology** | 32 (100.0) | 30 (93.75) | 29 (90.62) | 32 (100.0) | 28 (87.5) | 28 (87.5) | 23 (71.88) | 31 (96.88) |
| **Orthopedics** | 30 (93.75) | 26 (81.25) | 27 (84.38) | 28 (87.5) | 26 (81.25) | 22 (68.75) | 28 (87.5) | 30 (93.75) |
| **Pediatrics** | 26 (81.25) | 19 (59.38) | 13 (40.62) | 15 (46.88) | 23 (71.88) | 16 (50.0) | 12 (37.5) | 20 (62.5) |
| **Psychiatry** | 31 (96.88) | 29 (90.62) | 29 (90.62) | 28 (87.5) | 26 (81.25) | 27 (84.38) | 28 (87.5) | 26 (81.25) |
| **Total satisfaction** | 119 (92.97) | 104 (81.25) | 98 (76.56) | 103 (80.47) | 103 (80.47) | 93 (72.66) | 91 (71.09) | 107 (83.59) |
| **Search type** | | | | | | | | |
| **Diagnosis** | 28 (87.5) | 23 (71.875) | 23 (71.875) | 21 (65.625) | 21 (65.625) | 21 (65.625) | 22 (68.75) | 25 (78.125) |
| **General Knowledge** | 28 (87.5) | 25 (78.125) | 22 (68.75) | 25 (78.125) | 26 (81.25) | 21 (65.625) | 24 (75.0) | 24 (75.0) |
| **Management** | 32 (100.0) | 28 (87.5) | 26 (81.25) | 31 (96.875) | 28 (87.5) | 27 (84.375) | 23 (71.875) | 32 (100.0) |
| **Research** | 31 (96.875) | 27 (84.375) | 27 (84.375) | 26 (81.25) | 28 (87.5) | 24 (75.0) | 22 (68.75) | 26 (81.25) |
| **Total satisfaction** | 119 (92.969) | 103 (80.469) | 98 (76.562) | 103 (80.469) | 103 (80.469) | 93 (72.656) | 91 (71.094) | 107 (83.594) |

**Supplementary Table S12.** Expert satisfaction with model responses across specialties (Kruskal–Wallis and post hoc comparisons)

| Pos Hoc Dwass-Steel-Critchlow-Fligner | | p |
|---|---|---|
| Gynecology | Orthopedics | 0.133 |
| Gynecology | Pediatrics | <.001 |
| Gynecology | Psychiatry | 0.574 |

| | | |
|---|---|---|
| Orthopedics | Pediatrics | <.001 |
| Orthopedics | Psychiatry | 0.808 |
| Pediatrics | Psychiatry | <.001 |

Kruskal-Wallis p < 0.001 Post hoc tests indicated that Pediatrics differed significantly from Gynecology (p<0.001), Orthopedics (p<0.001), and Psychiatry (p<0.001). No significant differences were observed among Gynecology, Orthopedics, and Psychiatry (all p>0.05).

**Supplementary Table S13.** Expert satisfaction with model responses by type of question type and agent (Kruskal–Wallis and post hoc comparisons)

| Pos Hoc Dwass-Steel-CritchlowFligner | | p |
|---|---|---|
| Diagnosis | General Knowledge | 0.543 |
| Diagnosis | Management | 0.03 |
| Diagnosis | Research | 0.118 |
| General Knowledge | Management | 0.082 |
| General Knowledge | Research | 0.385 |
| Management | Research | 0.479 |

Note: Kruskal–Wallis test indicated overall differences across question types (p=0.007). Post hoc tests showed that Diagnosis differed significantly from Management (p=0.03); all other pairwise comparisons were not significant (p>0.05).

**Supplementary Table S14**. Specialist satisfaction disaggregated by evaluation criterion and specialty.

| | **Correct information n (%)** | | | **Agrees with consensus n (%)** | | | **Non biased n (%)** | | | **No recommendation against standard of care n (%)** | | | **Up to date n (%)** | | | **No risk for patient n (%)** | | | **Real reference sources n (%)** | | | **Context-awareness n (%)** | | | |
|---|---|---|---|---|---|---|---|---|---|---|---|---|---|---|---|---|---|---|---|---|---|---|---|---|---|
| | **Satisfaction n=118 (92.188)** | | | **Satisfaction n=122 (95.312)** | | | **Satisfaction n=107 (83.594)** | | | **Satisfaction n=88 (68.750)** | | | **Satisfaction n=111 (86.719)** | | | **Satisfaction n=102 (79.688)** | | | **Satisfaction n=72 (56.250)** | | | **Satisfaction n=97 (75.781)** | | | |
| **Specialty** | Somewhat satisfied n (%) | Very satisfied n (%) | Median (RIQ) | Somewhat satisfied n (%) | Very satisfied n (%) | Median (RIQ) | Somewhat satisfied n (%) | Very satisfied n (%) | Median (RIQ) | Somewhat satisfied n (%) | Very satisfied n (%) | Median (RIQ) | Somewhat satisfied n (%) | Very satisfied n (%) | Median (RIQ) | Somewhat satisfied n (%) | Very satisfied n (%) | Median (RIQ) | Somewhat satisfied n (%) | Very satisfied n (%) | Median (RIQ) | Somewhat satisfied n (%) | Very satisfied n (%) | Median (RIQ) | Total Satisfaction N (%) |
| **Gynecology** | 14 (43.8) | 15 (46.9) | 5 (1) | 11 (34.4) | 20 (62.5) | 5 (1) | 8 (25.0) | 23 (71.9) | 5 (1) | 5 (15.6) | 27 (84.4) | 5 (1) | 17 (53.1) | 8 (25.0) | 4 (1) | 7 (21.9) | 25 (78.1) | 5 (1) | 8 (25.0) | 13 (40.6) | 5 (1) | 0 (0.0) | 32 (100.0) | 5 (0) | 233 (91.0) |

| | | | | | | | | | | | | | | | | | | | | | | | | | |
|---|---|---|---|---|---|---|---|---|---|---|---|---|---|---|---|---|---|---|---|---|---|---|---|---|---|
| **Orthopedics** | 11 (34.4) | 21 (65.6) | 5 (1) | 9 (28.1) | 23 (71.9) | 5 (1) | 7 (21.9) | 21 (65.6) | 5 (1) | 2 (6.2) | 22 (68.8) | 5 (1) | 3 (9.4) | 29 (90.6) | 5 (1) | 0 (0.0) | 28 (87.5) | 5 (0) | 7 (21.9) | 9 (28.1) | 5 (1) | 4 (12.5) | 21 (65.6) | 5 (1) | 217 (84.8) |
| **Pediatrics** | 4 (12.5) | 23 (71.9) | 5 (1) | 3 (9.4) | 25 (78.1) | 5 (1) | 3 (9.4) | 14 (43.8) | 5 (1) | 0 (0.0) | 10 (31.2) | 5 (0) | 9 (28.1) | 13 (40.6) | 5 (1) | 2 (6.2) | 10 (31.2) | 5 (1) | 3 (9.4) | 13 (40.6) | 5 (1) | 3 (9.4) | 8 (25.0) | 5 (1) | 143 (55.9) |
| **Psychiatry** | 11 (34.4) | 19 (59.4) | 5 (1) | 5 (15.6) | 26 (81.2) | 5 (1) | 0 (0.0) | 31 (96.9) | 5 (0) | 1 (3.1) | 21 (65.6) | 5 (1) | 3 (9.4) | 29 (90.6) | 5 (1) | 2 (6.2) | 28 (87.5) | 5 (1) | 5 (15.6) | 14 (43.8) | 5 (1) | 0 (0.0) | 29 (90.6) | 5 (0) | 224 (87.5) |
